# Direct influence of BMPR2 mutations on cytokine patterns and biomarker effectiveness in pulmonary arterial hypertension

**DOI:** 10.1101/2021.05.05.21253970

**Authors:** Max Schwiening, Emilia M Swietlik, Divya Pandya, Keith Burling, Peter Barker, Carmen Treacy, Susana Abreu, S. John Wort, Joanna Pepke-Zaba, Stefan Graf, Stefan J Marciniak, Nicholas Morrell, Elaine Soon, members of the UK National Cohort Study of Idiopathic and Heritable PAH

**Affiliations:** Cambridge Institute for Medical Research, University of Cambridge, Keith Peters Building, Hills Rd, Cambridge CB2 0XY, UK; Department of Respiratory Medicine, Level 5, Box 157, Cambridge University Hospitals NHS Foundation Trust, Hills Road, Cambridge CB2 0QQ, UK; NIHR Cambridge BRC Core Biochemical Assay Laboratory (CBAL), Box 232, Cambridge University Hospitals NHS Foundation Trust, Hills Road, Cambridge CB2 0QQ, UK; National Heart and Lung Institute, Imperial College London, Guy Scadding Building, Dovehouse St, Chelsea, London SW3 6LY; Royal Papworth Hospital NHS Foundation Trust, Papworth Rd, Trumpington, Cambridge CB2 0AY; Department of Haematology, University of Cambridge, Cambridge Biomedical Campus, Cambridge CB2 0PT, UK; NIHR BioResource for Translational Research, Cambridge Biomedical Campus, Cambridge CB2 0QQ, UK

## Abstract

**Background:** Pulmonary arterial hypertension (PAH) covers a range of life-limiting illnesses characterized by increased pulmonary arterial pressures leading to right heart failure and death, if untreated. 15-25% of patients have genetic mutations, the most common affecting bone morphogenetic protein receptor type 2 (*BMPR2)*. The aim was to define an inflammatory cytokine profile in *BMPR2*-mutation positive patients and analyze their influence on survival.

**Methods:** Levels of cytokines were measured in plasma samples from *BMPR2*-mutation positive patients (*BMPR2mut*, n=54), patients without any driving mutations (n=54), and healthy controls (n=56) recruited from the United Kingdom cohort.

**Findings:** *BMPR2-*mutation positive patients and patients without mutations had high levels of interleukin-6, interleukin-8, tumor necrosis factor-α, and vascular endothelial growth factor-A compared to controls. Only *BMPR2-*mutation carrying patients had higher G-CSF levels compared to controls. VEGF-A levels were substantially higher in patients without mutations compared to the *BMPR2mut* group. Interleukin-6 was a significant discriminator for mortality in the *BMPR2mut* cohort (cumulative survival with interleukin-6≥1.6pg/ml at 3 years was 65% compared to 96% with interleukin-6<1.6pg/ml, *P*=0·0013). N-Terminal pro-B-Type natriuretic peptide levels did not discriminate for survival in our *BMPR2mut* cohort (cumulative survival for patients with an NT-proBNP>130ng/ml at 3 years was 76% compared to 84% for patients with an NT-proBNP≤130ng/ml, *P*=0·37). NT-proBNP outperformed interleukin-6 in PAH without mutations.

**Interpretation:** *BMPR2*-mutation positivity has a direct impact not only on inflammatory profiles but also on effectiveness of prognostic biomarkers. In our *BMPR2*-mutation positive cohort IL-6 was the strongest prognostic biomarker and NT-proBNP failed to discriminate for survival.

**Key messages:** *What is the key question?:* Do pulmonary arterial hypertension patients who are BMPR2-mutation positive have a different cytokine signature than PAH patients without mutations?

*What is the bottom line?:* BMPR2-mutation positive and PAH patients without mutations display different patterns of cytokine elevation and these cytokines differ in the way they influence transplant-free survival.

*Why read on?:* In our cohort of BMPR2-mutation positive patients, IL-6 is the best prognostic biomarker while NT-proBNP failed to discriminate for survival – this implies that prognostic biomarkers and by inference treatments could be genotype-specific.

**Graphical Abstract:** 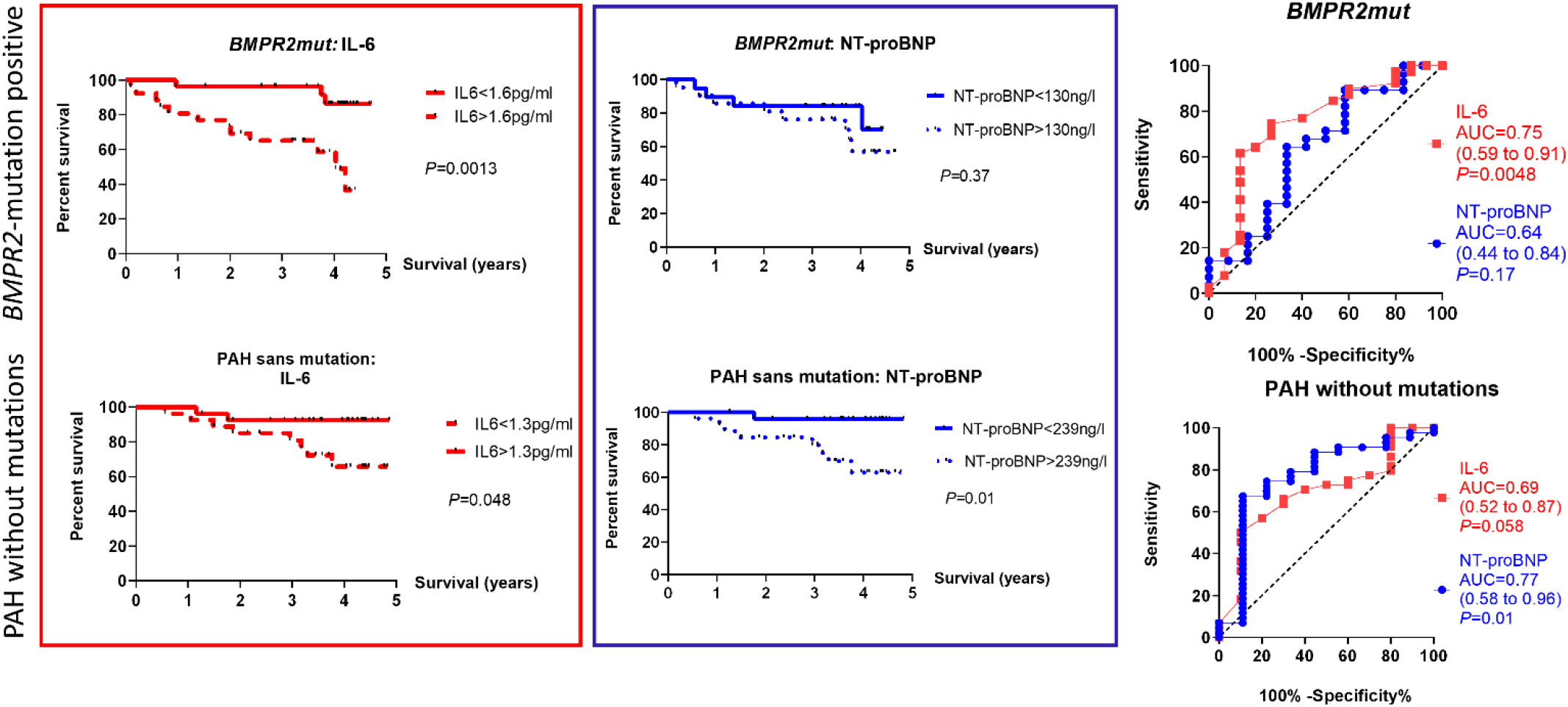

*TAKE HOME MESSAGE:* BMPR2-mutation positive patients have different inflammatory and growth factor profiles compared to PAH patients without mutations. Interleukin-6 is an effective biomarker for transplant-free survival in our cohort of *BMPR2*-mutation positive patients while NT-proBNP is ineffective. Conversely, NT-proBNP appears to be a more effective biomarker for pulmonary arterial patients without any mutations.

## Introduction

Pulmonary arterial hypertension (PAH) covers a range of life-limiting illnesses characterized by increased mean pulmonary arterial pressures, which if left untreated, lead to right heart failure and eventually death. Idiopathic PAH or PAH without any identifiable cause is due to remodeling of the small-to-medium sized pulmonary arteries which obstruct pulmonary blood flow and lead to right heart strain and eventually failure. The median survival for idiopathic PAH was 2.5 years(1). Over the last 30 years median survival has improved to 5.3 years(2) with the use of targeted therapies that mostly act as vasodilators of the pulmonary vascular bed. With the advent of modern genotyping techniques, it is now possible to subdivide the broad group of ‘idiopathic PAH’ into various forms of heritable PAH, characterized by mutations in specific genes, the most common affecting bone morphogenetic protein receptor type II (*BMPRII*)(3, 4), and idiopathic PAH without any known driver mutations. *BMPR2*-mutation positive patients are generally younger and have worse hemodynamic indices on diagnosis(5). It is estimated that the prevalence of *BMPR2* mutation in PAH is somewhere in the region of 10-26%(3-5). This then begs the question as to whether the mutation-positive groups have different underlying pathogenetic mechanisms and require different biomarkers and treatments, analogous to how EGFR-mutation positive non-small cell lung cancer patients respond to tyrosine kinase inhibition while the majority of NSCLC patients do not.

Multiple pre-clinical studies support the role of pro-inflammatory mechanisms in the pathogenesis of PAH(6-8), and we have previously shown dysregulation of a broad range of inflammatory cytokines in PAH which have a direct impact on patient survival(9). We wondered if the broad pro-inflammatory phenotype seen in idiopathic PAH would be different in the heritable PAH subgroup. To answer questions like these, the National Cohort Study of Idiopathic and Heritable PAH (http://www.ipahcohort.com/) was set up in 2014 and designed to recruit and follow-up every patient in the UK with a diagnosis of idiopathic or heritable PAH; and also to undertake whole genome sequencing as part of the NIHR BioResource for Rare Diseases(10, 11). This has given us an opportunity to examine the largest cohort of *BMPR2*-mutation positive patients to be characterized from an inflammatory perspective so far; and the opportunity to compare them with a contemporaneous group of PAH patients without any driving mutations.

## Methods

### Study subjects

All newly diagnosed patients with a confirmed diagnosis of idiopathic or heritable PAH were prospectively recruited at the national pulmonary hypertension centres in the UK from 2014 onwards under the aegis of the National Cohort Study of Idiopathic and Heritable PAH. Patients with a pre-existing diagnosis were also approached and asked for consent to be included. Idiopathic PAH was defined as a mean pulmonary artery pressure of ≥25mmHg at rest with a pulmonary capillary wedge pressure of ≤15mmHg with no underlying cause for PAH(12). Heritable PAH was defined as familial PAH (where 2 or more family members are affected) or where a pathogenic mutation in one of the known genes associated with PAH has been identified(13). This cohort was then characterized in the following manner:

a. Demographics including age at diagnosis, gender, and PAH centre,
b. Hemodynamic measurements from the diagnostic right heart catheterization,
c. Survival status at 31/12/2018 and survival time from sampling for cytokines to either death or transplantation,
d. N-terminal pro-B type natriuretic peptide (NT-proBNP) levels for each patient throughout the follow-up period,
e. the presence or absence of any known mutation predisposing to the development of PAH,
f. for *BMPR2*-mutation positive patients, the location and type of mutation.

Data harvesting was performed by a team of researchers blinded to the results of the cytokine analyses. A random sample of the extracted data was manually checked by separate researchers against the original database to ensure accuracy.

The National Cohort Study of Idiopathic and Heritable PAH also recruited fifty-six, gender-matched healthy controls. The baseline demographic data and medical history for these people were recorded in the same data capture system along with data collected for the cases. The study complies with the Declaration of Helsinki and was approved by the local research ethics committee (REC REF: 13/EE/0325 and 13/EE/0203). All participants gave informed written consent.

### Genetic analysis

All patients systematically underwent next-generation paired-end whole-genome sequencing using Illumina HiSeq2500 and HiSeq X (Illumina Inc, USA). Please see Supplement for details.

### Cytokine measurement

Blood sampling for cytokines and growth factors was performed at baseline in each subject during the initial study visit and then afterwards at timepoints which coincided with clinic visits. Survival time was calculated individually as the period from blood sampling or clinical measurement to either death or transplantation. The median follow-up time was 3.70 (2.85-4.2) years. Venous blood samples were collected in plasma-citrate tubes, mixed by inversion 3-4 times, and centrifuged within 40-60 minutes at 1,500*g* for 15 minutes. These samples were aliquoted and stored at -80°C until analyzed. Cytokines and growth factors were measured using a custom multiplex assay manufactured by Meso Scale Discovery (MSD; Gaithersburg, MD) and performed to manufacturer’s protocols (https://www.mesoscale.com/en/products/u-plex-immuno-oncology-group-1-human-111-plex-k15342k/).

### Statistics

Data was tested for normality using the Kolmogorov-Smirnov method. The Mann-Whitney test was used for comparing non-parametric data and the unpaired t-test was used for parametric data. The Spearman test was performed for intergroup correlations. Median (interquartile range) are used in text, graphs, and tables for cytokine levels (which are largely non-parametric) and means (± standard deviations) are used for parametric data. To control the false discovery rate due to testing for multiple cytokines, we employed the Benjamini-Hochberg procedure(14) with a false discovery rate of 0.1. Kaplan-Meier curves were constructed using the median value for each cytokine or clinical parameter as a cut-off. A composite endpoint of death or transplant was used as a defining event for Kaplan-Meier analysis. Differences between Kaplan-Meier curves were analyzed using the log-rank test. *P*-values of <0.05 were considered statistically significant. Statistical analyses were performed using GraphPad Prism (v8.0.0 for Windows, GraphPad Software, California USA, www.graphpad.com), and the Handbook of Biological Statistics (http://www.biostathandbook.com/#print).

## Results

### Subject demographics

Table 1 shows the demographics for patients and controls. Controls were younger than all patients (mean age of controls being 36.6±12.5 years *versus* 43.7±12.8 years for all patients, *P*<0.01). Patients with *BMPR2* mutations (n=54) were younger than patients without mutations (n=54; 41.3±13.4 years *versus* 46.1±11.8 years, *P*<0.05) but had a higher mean pulmonary artery pressure (60.8±12.3 *versus* 52.7±8.4 mmHg, *P*<0.01) and a lower cardiac index on diagnosis (1.99±0.60 *versus* 2.27±0.70 L/min/m^2^, *P<*0.05). There were no differences in gender distribution between controls, *BMPR2*-mutation carrying patients and PAH patients without mutations.

**Table 1:** Baseline characteristics of all PAH patients.

| Demographic | Controls | PAH |  |  |
| --- | --- | --- | --- | --- |
|  | (healthy volunteers) | Total | BMPR2mut | PAH sans mutation |
| Number, n | 56 | 108 | 54 | 54 |
| Age (years) | 36.63 (12.50) | 43.72 (12.76)<sup>††</sup> | 41.25 (13.38)* | 46.10 (11.75)<sup>††</sup> |
| Male: Female | 1: 2.11 | 1: 2.50 | 1 : 1.95 | 1: 3.31 |
| mean PAP,<br>mmHg | N/A | 56.67 (11.20) | 60.76 (12.33)** | 52.73 (8.35) |
| CI, L/min/m2 | N/A | 2.14 (0.67) | 1.99 (0.60)* | 2.27 (0.70) |

\**P*<0.05, \*\**P*<0.01 compared to PAH without mutation
<sup>†</sup> *P*<0.01, <sup>††</sup> *P*<0.01 compared to controls.

### Analysis of cytokine and growth factor profiles

IL-6, IL-8, IL-10, TNF-α and VEGF-A were significantly elevated in all PAH patients *versus* controls (Table 2). *BMPR2-*mutation carrying patients have a different profile of cytokine and growth factors compared to PAH patients without mutations (Figure 1 and Table 2). Both *BMPR2-*mutation carrying patients and PAH patients had high levels of IL-6, IL-8 and TNF-α compared to controls. For example, the median level for IL-6 was 0.70 (0.60-0.90) pg/ml in controls compared to 1.60 (1.10-2.10) pg/ml for *BMPR2*-mutation positive patients and 1.35 (0.90-2.23) pg/ml in PAH patients without mutations (*P*<0.0001 for both). However only PAH patients without mutations had higher IL-10 levels compared to controls while only the *BMPR2-*mutation carrying patients had higher G-CSF levels compared to controls.Interestingly, PAH patients without mutations had much higher VEGF-A levels (48.50 [33.25-93.75] pg/ml) compared to *BMPR2-*mutation carrying patients (35.00 [26.00-54.00] pg/ml; *P*=0.0008).

**Table 2:** Plasma cytokine and growth factor levels in PAH patient groups versus healthy controls.

| Cytokine<br>(pg/ml) | Controls<br>n=56 | All PAH<br>n=108 | BMPR2mut<br>n=54 | PAH sans mutation<br>n=54 |
| --- | --- | --- | --- | --- |
| IL-6 | 0.70 (0.60-0.90) | 1.40 (1.00-2.10)<sup>***, ‡</sup> | 1.60 (1.10-2.10)<sup>***, ‡</sup> | 1.35 (0.90-2.23)<sup>***, ‡</sup> |
| IL-8 | 2.90 (2.50-3.60) | 3.70 (2.70-6.20)<sup>** , ‡</sup> | 3.60 (2.90-5.50)<sup>** , ‡</sup> | 3.85 (2.53 - 6.58)<sup>* , ‡</sup> |
| IL-10 | 0.30 (0.30-0.40) | 0.40 (0.30-0.50)<sup>* , ‡</sup> | 0.35 (0.30-0.50) | 0.40 (0.30-0.50)<sup>* , ‡</sup> |
| TNF-α | 1.90 (0.50-2.28) | 2.60 (2.10-3.20)<sup>*** , ‡</sup> | 2.50 (2.10-2.90)<sup>***, ‡</sup> | 3.00 (2.00-3.80)<sup>***, ‡</sup> |
| VEGF-A | 30.00 (25.00-38.00) | 40.00 (29.25-71.75)<sup>*** , ‡</sup> | 35.00 (26.00-54.00)<sup>†, ‡</sup> | 48.50 (33.25-93.75)<sup>*** , ‡</sup> |
| G-CSF | 12.00 (10.10-15.10) | 13.10 (11.30-17.00) | 13.35 (11.53-17.38)<sup>* , ‡</sup> | 12.65 (11.10-16.60) |

All values are expressed as medians (interquartile ranges).
\* $P < 0.05$ , \*\* $P < 0.01$ , \*\*\* $P < 0.001$ compared to controls using Mann-Whitney testing.
<sup>†</sup> $P < 0.05$ compared to PAH without mutations using Mann-Whitney testing.
<sup>‡</sup> $P$ reaches significance as determined by Benjamini-Hochberg testing with a false discovery rate of 0.1

**Figure 1.**
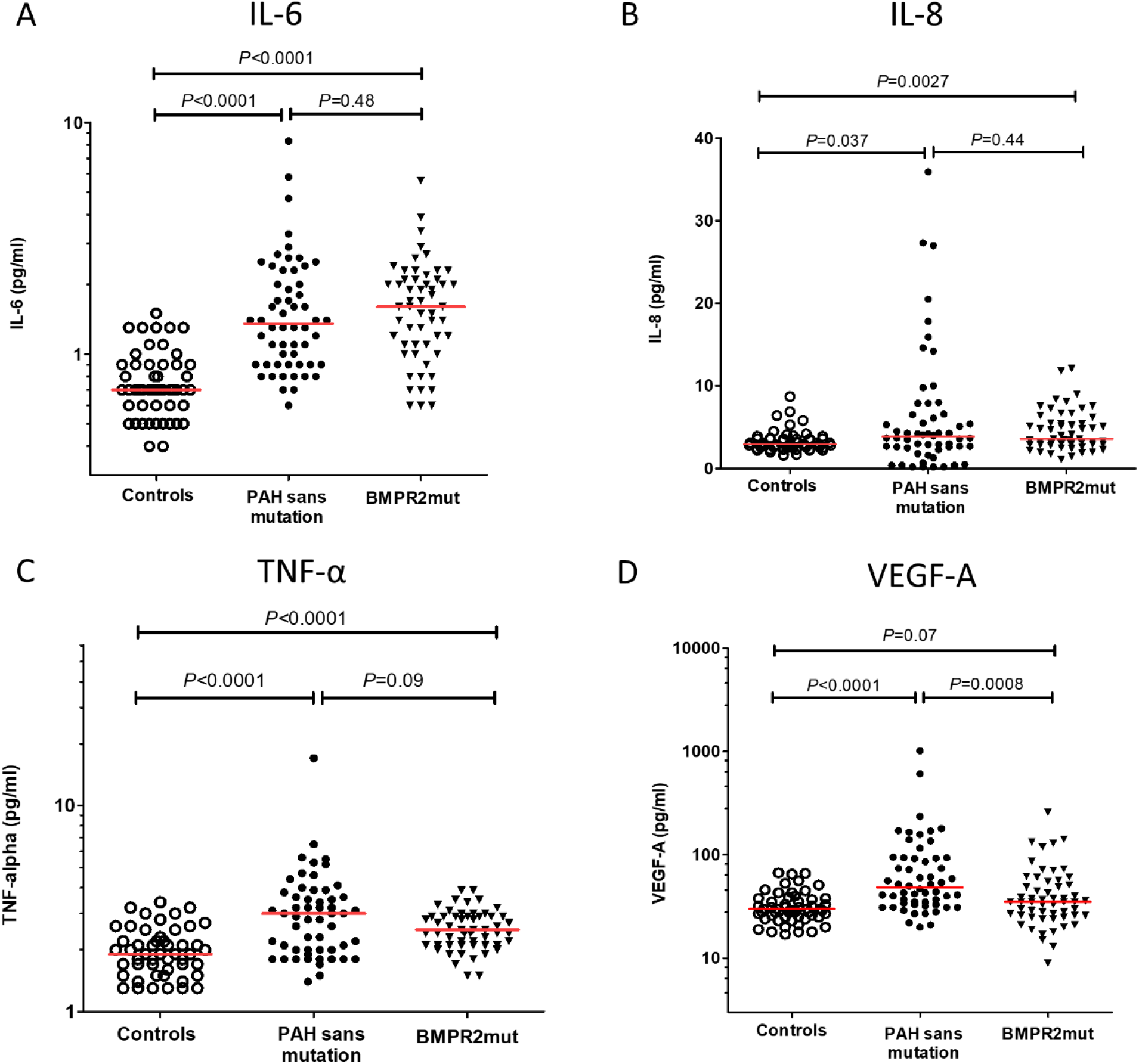
Graphs showing the distributions and medians of plasma cytokines and growth factors in *BMPR2-*mutation carrying patients (*BMPR2mut*) and PAH patients without any known mutations (PAH sans mutation). Graphs show (A) IL-6, (B) IL-8, (C) TNF-α, and (D) VEGF-A. Note that axes are logarithmic for (A), (C) and (D). Statistical significance was determined using the Mann-Whitney test. A single value of IL-8 is missing from (B) for the *BMPR2mut* cohort due to a technical issue with the assay.

### Correlations between different cytokines

Spearman correlations between different cytokines are shown in Table 3. The overwhelming majority of significant correlations occur in the PAH without mutations group. For example, TNF-α is significantly correlated with IL-10 (r=0.49), IL-6 (r=0.42), VEGF-A (r=0.46) and IL-8 (r=0.35) [P<0.01 in all cases] in PAH without mutations. The only highly significant correlation in the *BMPR2*-mutation carrying group is between TNF-α and IL-10 (r=0.46, *P*<0.001).

**Table 3:** Analysis of correlations between significantly elevated cytokines in PAH patient groups.

| IL-6 | IL-8 | IL-10 | TNF-α | VEGF-A | G-CSF | Cytokine/GF |
| --- | --- | --- | --- | --- | --- | --- |
| N/A | All PAH: r= 0.16<br>PAH sans mut: r= 0.21<br>BMPR2mut: r= 0.12 | All PAH: r= 0.21*<br>PAH sans mut: r= 0.21<br>BMPR2mut: r= 0.12 | All PAH: r= 0.35**<br>PAH sans mut: r= 0.42**<br>BMPR2mut: r= 0.21 | All PAH: r= 0.06<br>PAH sans mut: r= 0.033<br>BMPR2mut: r= 0.21 | All PAH: r= 0.12<br>PAH sans mut: r= 0.093<br>BMPR2mut: r= 0.13 | IL-6 |
|  | N/A | All PAH: r= 0.17<br>PAH sans mut: r= 0.15<br>BMPR2mut: r= 0.16 | All PAH: r= 0.33***<br>PAH sans mut: r= 0.35**<br>BMPR2mut: r= 0.25 | All PAH: r= 0.37**<br>PAH sans mut: r= 0.61***<br>BMPR2mut: r= 0.15 | All PAH: r= 0.0077<br>PAH sans mut: r= 0.093<br>BMPR2mut: r= 0.13 | IL-8 |
|  |  | N/A | All PAH: r= 0.48***<br>PAH sans mut: r= 0.49***<br>BMPR2mut: r= 0.46*** | All PAH: r= 0.057<br>PAH sans mut: r= 0.13<br>BMPR2mut: r= -0.075 | All PAH: r= 0.19*<br>PAH sans mut: r= 0.24<br>BMPR2mut: r= 0.16 | IL-10 |
| | | | N/A | All PAH: r= 0.35***<br>PAH sans mut: r= 0.46***<br>BMPR2mut: r= 0.13 | All PAH: r= 0.23*<br>PAH sans mut: r= 0.24<br>BMPR2mut: r= 0.30* | TNF-α |
|  |  |  |  | N/A | All PAH: r= 0.047<br>PAH sans mut: r= 0.83<br>BMPR2mut: r= 0.35 | VEGF-A |
|  |  |  |  |  | N/A | G-CSF |

5 Correlation coefficients are determined by the Spearman method.
\* $P<0.05$ , \*\* $P<0.01$ , \*\*\* $P<0.001$

### Survival analyses using cytokine levels as discriminators

We wondered if cytokine levels would be significant discriminators for transplant-free survival. The *BMPR2*-mutation carrying group had a median follow-up of 3.72 (2.79-4.19) years and 15 events were recorded in 54 patients. The PAH without mutation group had a median follow-up of 3.58 (2.93-4.36 years, *P*=0.64) and 10 events were recorded in 54 patients. We performed Kaplan-Meier analyses using the medians and tertiles of the cytokines (Figure 2, Supplementary Fig.1). For *BMPR2*-mutation carrying patients, IL-6 was a truly significant predictor of mortality – the cumulative survival for patients with IL-6 of ≥1.6pg/ml at 3 years is 65% compared to 96% for patients with an IL-6 of <1.6pg/ml (log-rank *P*=0.0013). For PAH patients without mutations, IL-6 also proved to be a significant discriminator for survival but to a lesser degree - the cumulative survival for patients with an IL-6 ≥1.3pg/ml at 3 years is 81% compared to 92% for patients with an IL-6 <1.3pg/ml (log-rank *P*=0.048). However, for PAH patients without mutations, TNF-α and IL-8 levels had a significant impact on survival which was not seen in *BMPR2*-mutation carrying patients. The cumulative survival for PAH patients without mutations with an IL-8 >4.2pg/ml at 3 years is 79% compared to 93% for patients with an IL-8 ≤4.2pg/ml (log-rank *P*=0.02), while the cumulative survival for *BMPR2*-mutation positive patients with an IL-8 >3.6pg/ml at 3 years is 77% compared to 85% for patients with an IL-8 <3.6pg/ml (log-rank *P*=0.09). Supplementary Table 1 summarizes the cumulative survivals at 1-, 2-, 3- and 4-year periods for *BMPR2*-mutation positive and PAH without mutations.

**Figure 2.**
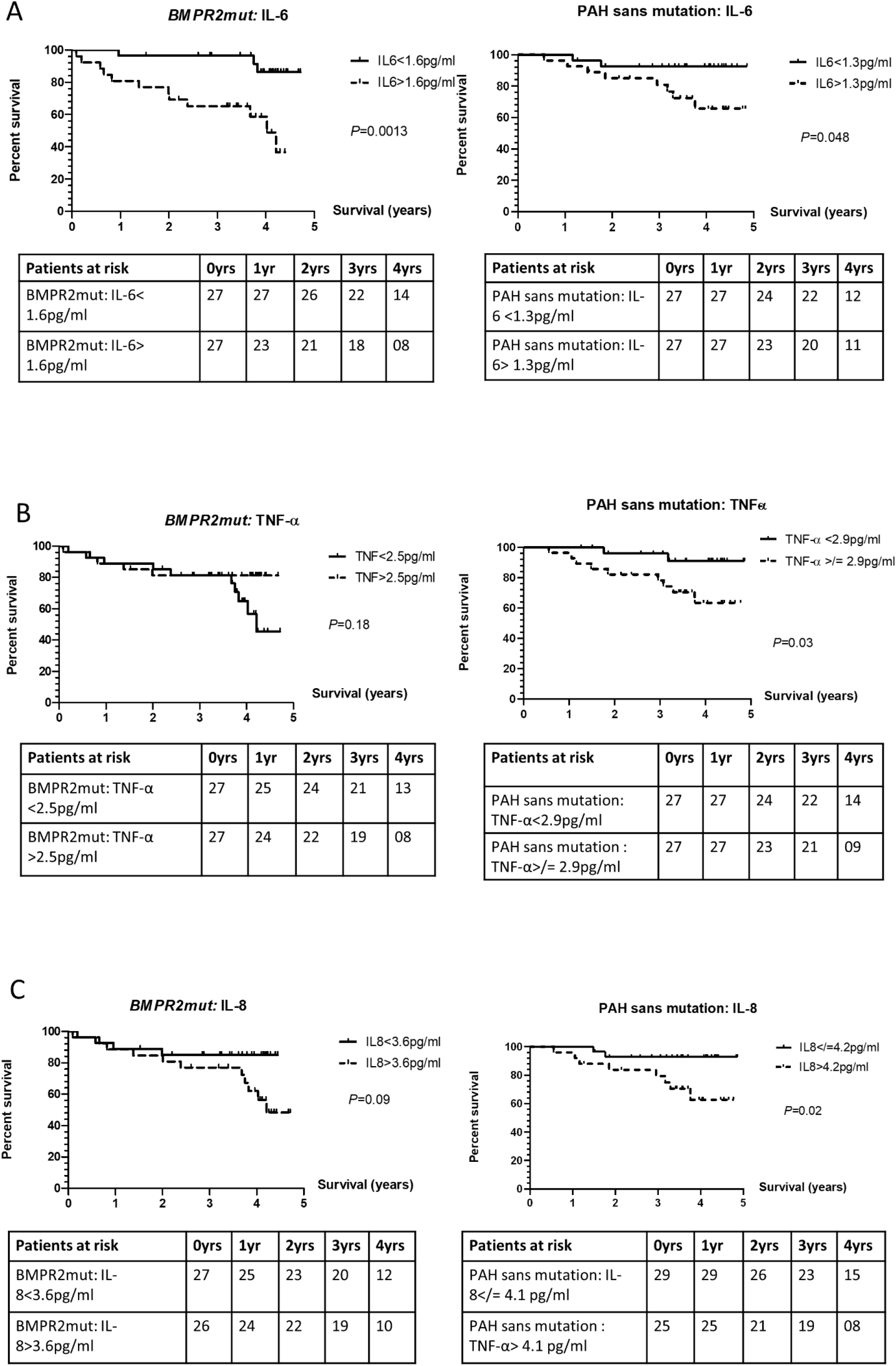
Kaplan-Meier analyses based on median levels of cytokines in *BMPR2-*mutation positive patients (*BMPR2mut*) and PAH patients without any known mutations (PAH sans mutation). Graphs show survival curves based on levels of (A) IL-6, (B) TNF-α, and (C) IL-8. Note that a single value of IL-8 is missing from (C) for the *BMPR2mut* cohort due to a technical issue with the assay.

### Survival analyses using hemodynamic parameters and NT-proBNP levels as discriminators

Cardiac index and right atrial pressure at diagnostic right heart catheterization have been previously established as important factors that predict mortality in PAH patients (1, 15, 16). This is true of our cohort of PAH patients without mutations. Interestingly, this does not hold in the *BMPR2*-mutation carrying patients (Figure 3). Furthermore, analyses of NT-proBNP levels measured concomitantly with cytokine levels reveal that NT-proBNP is a good marker of prognosis in PAH without mutations but not in *BMPR2*-mutation carrying patients (Figure 3A, Table 4). To ensure that this was not an error the survival analyses was redone using tertiles of NT-proBNP levels, and also using cut-off values that had been previously validated by the ERS(17) (with values of NT-proBNP<300ng/l being regarded as ‘low’ and values of NT-proBNP>1,400ng/l being regarded as ‘high’ – Supplementary Fig.2). No matter what cut-off value was used, NT-proBNP levels did not prove to be a discriminator for survival in *BMPR2*-mutation carrying patients. Matching analyses were done in the PAH patients without driving mutations (Supplementary Fig.2).

**Figure 3.**
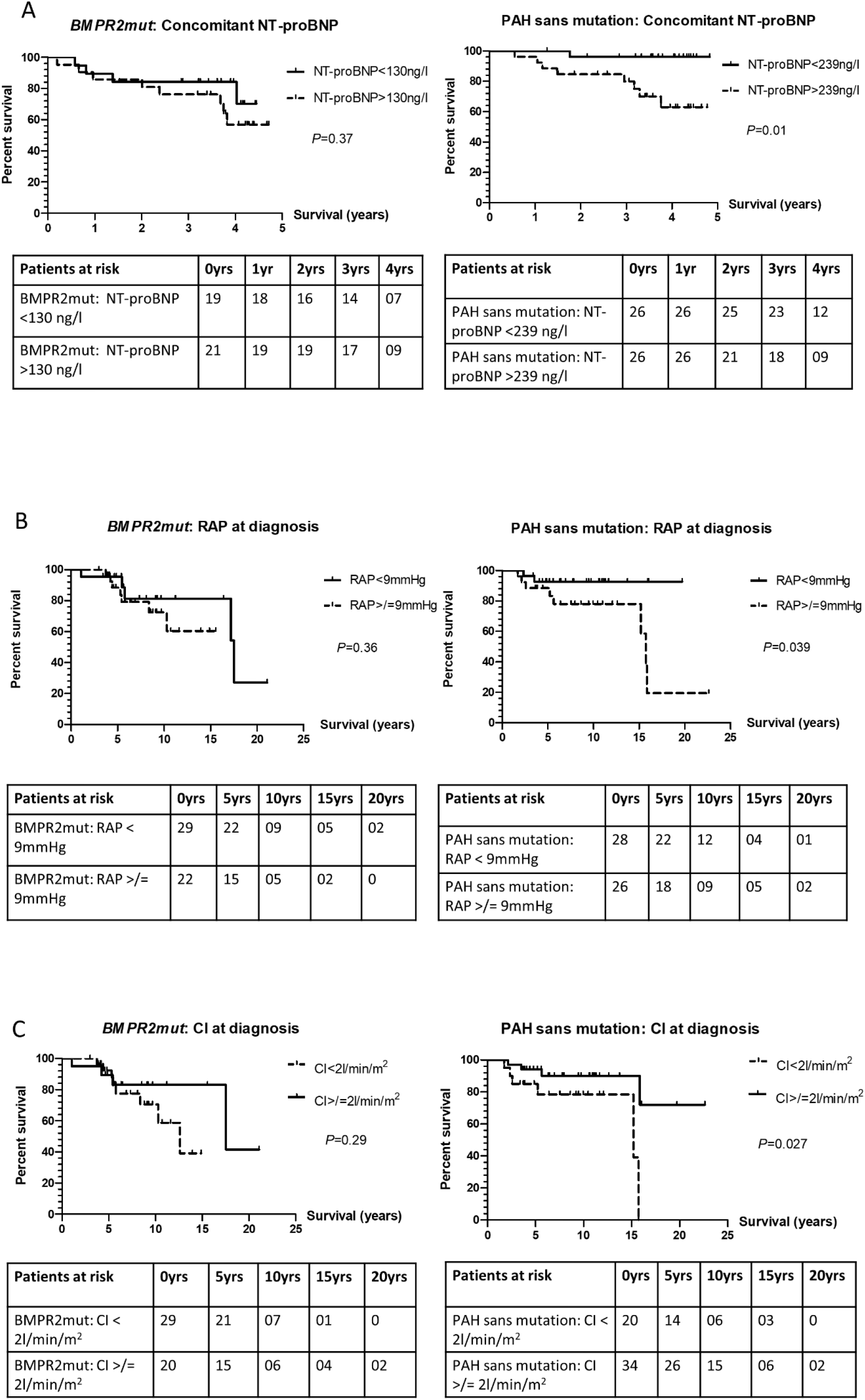
Kaplan-Meier analyses based on previously verified clinical parameters in *BMPR2-*mutation carrying patients (*BMPR2mut*) and PAH patients without any known mutations (PAH sans mutation). Graphs show survival curves based on (A) levels of N-terminal pro-B type natriuretic peptide [NT-proBNP] taken contemporaneously with the blood drawn for cytokine testing, (B) right atrial pressure [RAP] at diagnosis, and (C) cardiac index [CI] at diagnosis. Note that numbers do not add up to 54 as values are missing from some sections due to differences in follow-up between different centres (for NT-proBNP) and not all historical diagnoses having recorded full diagnostic RHC details on the database (*e*.*g*. a cardiac output only having been recorded at the diagnostic RHC). Where possible a cardiac index has been retrospectively calculated if the patient’s height and weight were available from the time of diagnosis.

**Table 4:**
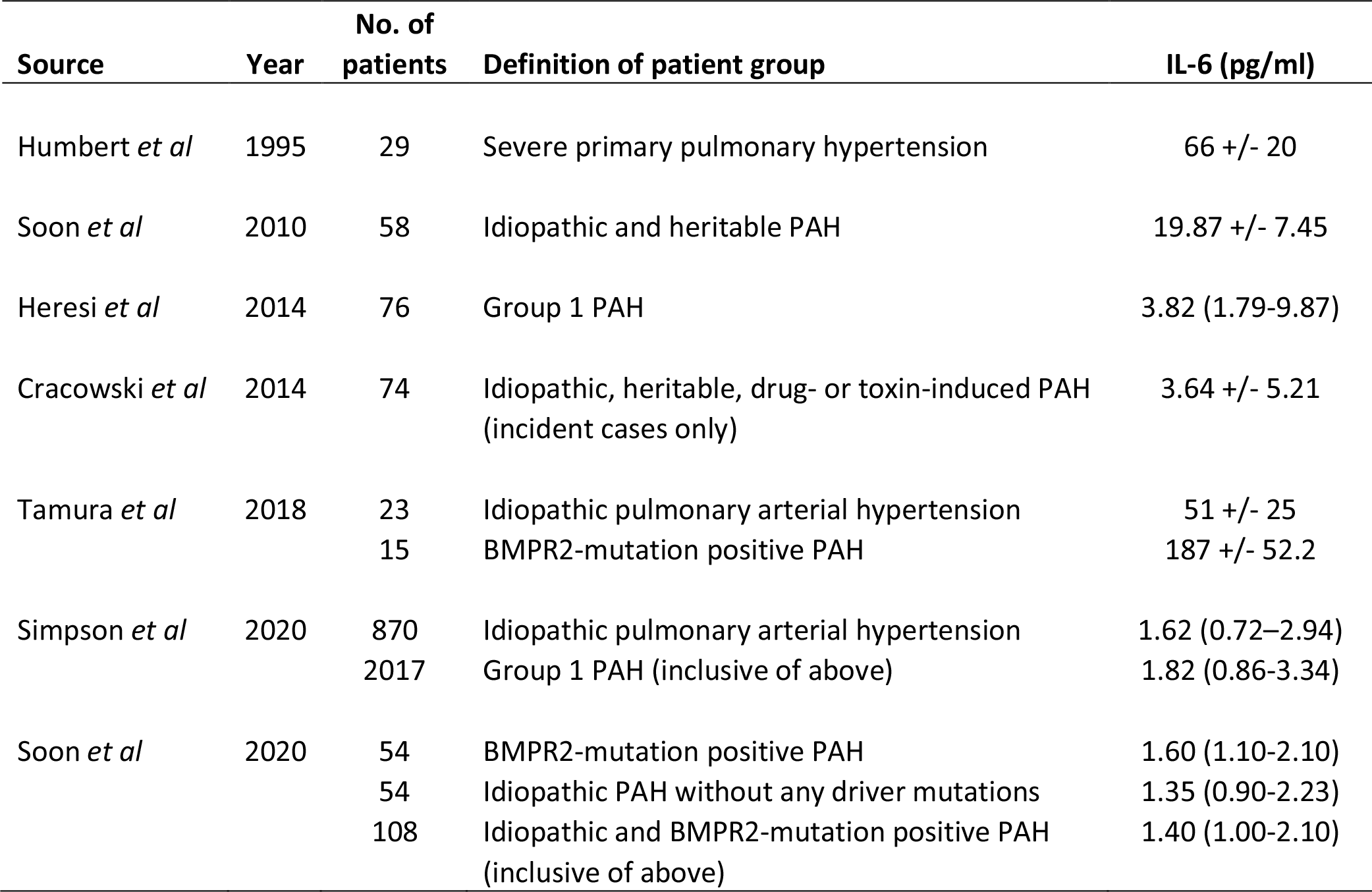
Representative IL-6 levels for idiopathic, heritable, group 1 PAH and BMPR2-mutation positive PAH patients from published data. Data are represented as either (mean +/− SD) or [median (interquartile range)].

### Survival analyses in BMPR2-mutation carrying patients as a sub-group

In this cohort, there was no survival disadvantage associated with the possession of a *BMPR2*-mutation compared to the group without any driving mutations (Figure 4A). Interestingly, patients with a *BMPR2* mutation affecting the kinase domain had worse survival compared to those with mutations in any other domain. For example, kinase domain mutation-carrying patients had a cumulative survival of 74% at 3 years in *versus* 89% in patients with mutations in all other domains (*P=0*.*02*, Figure 4B). However, we did not see a survival disadvantage associated with missense mutations (Figure 4C) nor with male gender (Figure 4D).

**Figure 4.**
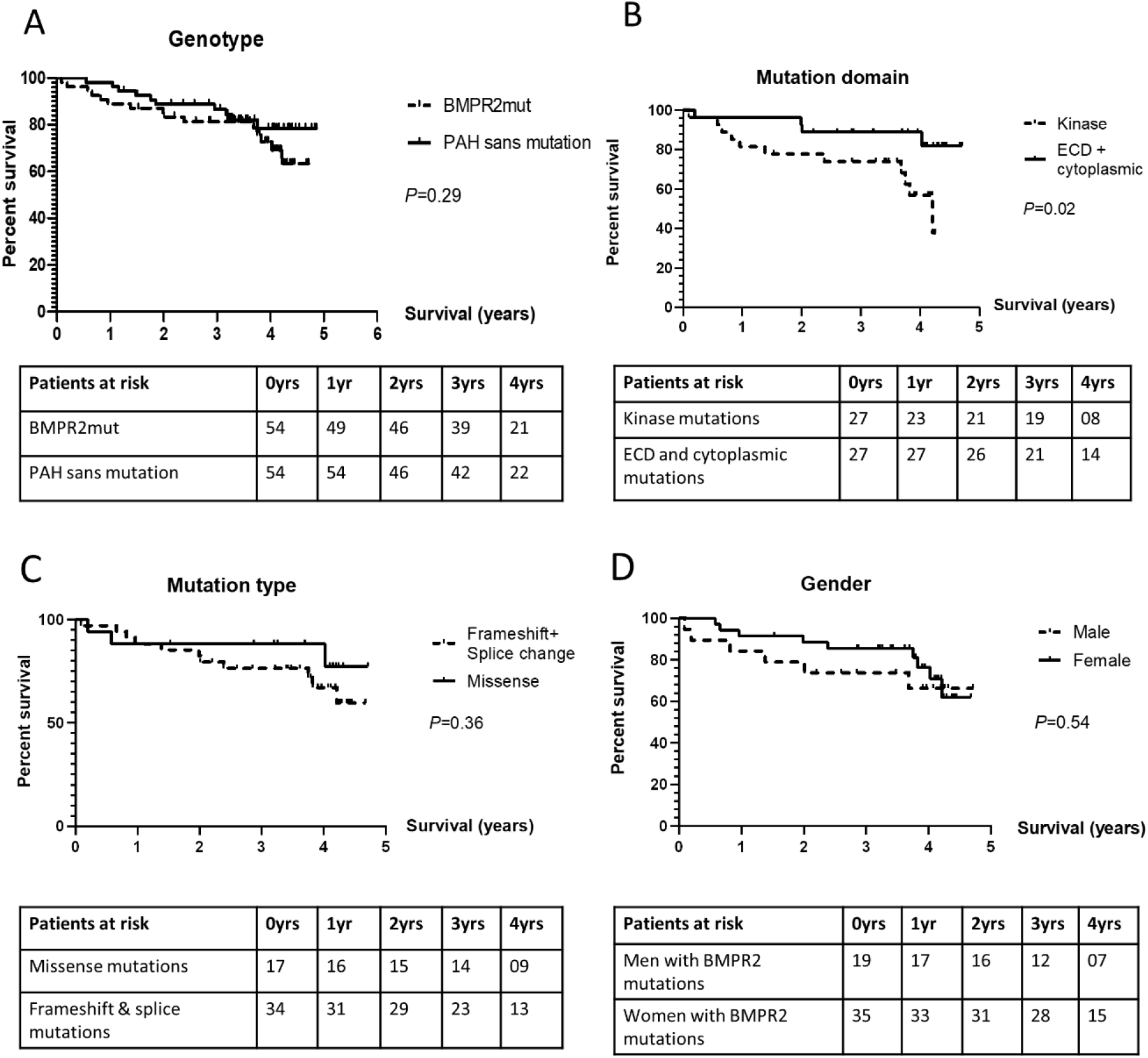
Kaplan-Meier analyses based on clinical and BMPR2-mutation parameters. Graphs show survival curves based on (A) the presence of a *BMPR2*-mutation *versus* no detectable mutation, (B) the domain in which the *BMPR2* mutation resides, (C) the type of *BMPR2* mutation, and (D) gender in *BMPR2-*mutation carrying patients (*BMPR2mut*). Note that for panel (C) 3 values are missing due to the presence of 2 stop-codon and 1 in-frame insertion which could not be easily classified.

### Longitudinal tracking of IL-6

We wondered if tracking the changes in plasma IL-6 levels would give any further prognostic information for these patients. To this end, we obtained follow-up plasma samples taken a year later. A year was chosen as the follow-up point to allow patients to receive and be stabilized on treatment regiments prior to sampling. 42 follow-up samples were available from the original cohort of 54 *BMPR2*-mutation positive patients; and 49 follow-up samples from the original cohort of 54 PAH patients without mutations. We were unable to find any differences in the change of IL-6 levels between patients who survived to the time of census and patients who died or were transplanted (Supplementary Fig.3A-B). When we examined the follow-up cohort more closely, we realized that 46.7% (7 of 15) the *BMPR2*-mutation positive patients who died/were transplanted were unable to contribute a follow-up sample as they died before the year had passed (Supplementary Fig.3C). Similarly, 4 of the 10 PAH patients without mutations who died/were transplanted did so before a year had passed and hence did not have a follow-up sample (Supplementary Fig.3D). Therefore, we did not proceed to any further analysis as the sample size had eroded significantly in the patient cohorts who died or had been transplanted.

## Discussion

We first reported the link between increased cytokine levels and worse survival in PAH patients in 2010(9). This study represents the chance to consolidate and extend that finding – we report results from the largest cohort of *BMPR2*-mutation positive patients to be characterized from an inflammatory perspective so far.

We confirm our previous finding that IL-6, IL-8 and TNF-α levels are elevated in both heritable and idiopathic PAH patients, as we first demonstrated in 2010. This has been further substantiated by Cracowski(18), Simpson(19) and Heresi(20). *BMPR2*-mutation carrying patients have a subtly different cytokine profile, with lower VEGF-A and IL-10 levels and higher G-CSF levels. The impact of cytokines on mortality is also different between the *BMPR2*-mutation positive group and PAH *sans* mutations. Only IL-6 appears to be an important predictor of mortality in the *BMPR2*-mutation carrying cohort; while IL-6, TNF-α and IL-8 remained significant discriminators in the PAH without driving mutations group.

This is the first direct comparison of IL-6 and NT-proBNP in BMPR2-mutation positive patients. IL-6 is a more sensitive discriminator for mortality in our BMPR2-mutation carrying patients than NT-proBNP (Fig.2A *versus* Fig.3A; Supplementary Fig.1A *versus* Supplementary Fig.2A and 2C). No matter what cut-offs were used for NT-proBNP (median levels in Fig. 3A, tertiles in Supplementary Fig. 2A and ERS/ESC definitions for ‘low’ and ‘high’ values of NT-proBNP(17) in Supplementary Fig. 2C) we were unable to find a level that served as a discriminator for mortality. This is markedly different in the PAH without driving mutations group (corresponding Kaplan-Meier curves are shown in Fig. 3B [median], Supplementary Fig.2B [tertiles] and Supplementary Fig. 2D [ERS values]) – all curves remain significant no matter which levels are used. This is also supported by the corresponding ROC curves (Graphical Abstract) – the area under curve (AUC) for IL-6 in *BMPR2*-mutation positive patients is 0.75 (*P*=0.0048) *versus* 0.64 for NT-proBNP (*P*=0.17). Conversely the AUC for IL-6 in PAH patients without mutations is 0.77 (*P*=0.01) *versus* 0.69 for IL-6 (*P*=0.058). Therefore, we posit that IL-6 is a better biomarker for prognosis in the *BMPR2*-mutation carrying patients while NT-proBNP remains superior in PAH without driving mutations.

The presence of *BMPR2*-mutation positive patients in different proportions in the overall PAH group might help to explain some of the differences seen in prior papers. For example, Cracowski reported that higher levels of IL-1α, IL-1β, TNF-α and IL-13 remained independently associated with mortality, while the impact of IL-6 was lost after adjusting for potential confounders such as age and right atrial pressure(18). On the other hand, Simpson reports that each log-unit increase of IL-6 was associated with a 31% higher risk of death(19). This may be accounted for if there were different proportions of BMPR2-mutation positive patients in the different study groups.

A potential difficulty in using IL-6 as a prognostic marker would be deciding what cut-off level to choose. Table 4 shows representative values from the papers that have measured IL-6 in idiopathic, heritable, and group 1 PAH patients. What leaps out is that levels of IL-6 have decreased markedly over time, with representative values of 66pg/ml in 1995(21) to 20pg/ml in 2010(9) to 1.4-1.62(19) pg/ml in 2020. This may be a result of more severe disease and/or less aggressive treatment in the earlier cohorts, different genetic backgrounds, or a combination thereof. It is likely that disease severity is a major factor as the mean cardiac index was 2.0 l/min/m^2^ in the 2010 cohort compared to 2.7 l/min/m^2^ and 2.14 l/min/m^2^ in the two 2020 cohorts. The exception to this trend is the cohort described by Tamura(22) – we posit that this may be due to illness severity as the 74% (17 of 23) IPAH patients and 93% (14 of 15) *BMPR2*-mutation positive patients were on 2 or more targeted treatments. We also note that the inclusion of different types of type I PAH (notably connective tissue disease associated PAH, CTD-PAH) has the potential to push the median levels for IL-6 upwards. For example, Simpson(19) reported median levels of 2.24 (1.09-4.33) pg/ml in CTD-PAH and 1.62 (0.72-2.94) pg/ml in IPAH. This implies that IL-6 levels are disease and genotype-specific. We suggest a prognostic cut-off of 1.6pg/ml as being useful in both *BMPR2*-positive and PAH without mutations (Supplementary Fig.4A-B) in our cohort and matching the results of the largest IPAH cohort available. This also takes into account that testing for *BMPR2* mutations may not be readily available. Clearly this requires prospective validation in an independent patient group.

What is also most interesting is that there appears to be a group within the PAH sans driving mutation cohort that is highly inflammatory in nature. Table 3 reveals that there are highly significant correlations (*P*<0.01) between levels of TNF-α, IL-6, IL-8, IL-10, and VEGF-A in PAH patients without driving mutations. No highly significant correlations between the various cytokines exist in *BMPR2*-positive PAH patients save for a single one between TNF-α and IL-10. This begs the question as to whether there is a subgroup within the PAH without mutations cohort whose driving force is inflammation. If such a group does exist, it seems reasonable to target this group with anti-inflammatory strategies. Conversely it appears that the main cytokine driver in *BMPR2*-positive PAH is IL-6. Strategies that target the IL-6 axis, such as tocilizumab, have proven successful in PAH associated with specific diseases including Castleman’s disease, MCTD and Still’s disease(23-26) and should be considered for trials targeting *BMPR2*-mutation positive disease.

This study raises the question of why there are differences in the inflammatory cytokine profiles in *BMPR2*-mutation positive PAH *versus* PAH without driving mutations. From the current published body of evidence we know that *BMPR2* mutations directly predispose to greater susceptibility to pro-inflammatory stimuli such as LPS(27) which results in higher levels of IL-6. We also know that TNF-α can directly the suppress BMPR-II signaling axis(28). Therefore, one can speculate that in the ‘inflammatory’ subset of PAH without mutations a trigger has led to the following sequence of events:

Inflammatory trigger > TNF-α production > suppression of BMPR-II signaling > greater production of IL-6 and IL-8 > pulmonary vascular remodeling….

This hypothesis explains the picture we see; *i*.*e*. the *BMPR2-*mutation positive cohort starts ‘a step ahead’ with constitutive genetic loss of BMPR-II and hence displays IL-6 as its main inflammatory driver; while in PAH without driving mutations both TNF-α and IL-6 are implicated in the underlying pathogenetic processes.

The strength of this study is that it is a multi-centre, national registry-based study which aims to collect data and samples prospectively and without bias from all patients in the UK. However, it does rely on the pre-existing clinical structure as registry visits are timed to co-incide with clinical visits and blood samples are taken at the same time as clinical samples to lessen the inconvenience and discomfort for patients. Thus, the follow-up differs between patients as patients who are sicker or not doing as well as expected on treatment will be seen more frequently than patients who are stable. A further strength is that the researchers who performed the cytokine measurements were blinded to the genotypes and clinical outcomes of the patients.

## Conclusions

We have identified distinct inflammatory and growth factor profiles in *BMPR2*-mutation positive PAH and PAH without driving mutations. IL-6 is the most discriminating biomarker for prognosis in our *BMPR2*-mutation positive PAH. NT-proBNP does not appear to be an effective biomarker for prognosis in this cohort of *BMPR2*-mutation positive PAH. NT-proBNP outperforms IL-6 in PAH without driving mutations. We postulate that BMPR2-mutation positive PAH may have a different driving inflammatory component and respond to different treatments compared to PAH patients without mutations.

## Data Availability

The data underlying this article will be shared on reasonable request to the corresponding author. The requester must have appropriate ethical permissions for this to happen.

## Acknowledgments

This work could not have been done without the British Heart Foundation/Medical Research Council (UK) National Cohort of Idiopathic and Heritable PAH. We thank the National Institute for Health Research and NHS Blood and Transplant, NIHR BioResource volunteers for their participation, and gratefully acknowledge NIHR BioResource centres, NHS Trusts, and staff for their contribution. We thank Ms. Tolulope Osunnuyi, Ms. Diana Hoult, Ms. Carol Dorling, and Ms. Robyn Murdoch for assisting with sample collection and Ms. Denise Hodgkins and Ms. Natalie Doughty for starting the PAH patient database. Most importantly special thanks to all our patients, families, and staff in the pulmonary hypertension centres in the UK who have selflessly donated their time, data, and tissue.

The views expressed are those of the authors and not necessarily those of the NHS, the NIHR or the Department of Health and Social Care.

## References

1. D’Alonzo GE, Barst RJ, Ayres SM, Bergofsky EH, Brundage BH, Detre KM, et al. Survival in patients with primary pulmonary hypertension. Results from a national prospective registry. Ann Intern Med. 1991;115(5):343–9.

2. Ling Y, Johnson MK, Kiely DG, Condliffe R, Elliot CA, Gibbs JS, et al. Changing demographics, epidemiology, and survival of incident pulmonary arterial hypertension: results from the pulmonary hypertension registry of the United Kingdom and Ireland. Am J Respir Crit Care Med. 2012;186(8):790–6.

3. Thomson JR, Machado RD, Pauciulo MW, Morgan NV, Humbert M, Elliott GC, et al. Sporadic primary pulmonary hypertension is associated with germline mutations of the gene encoding BMPR-II, a receptor member of the TGF-beta family. J Med Genet. 2000;37(10):741–5.

4. Lane KB, Machado RD, Pauciulo MW, Thomson JR, Phillips JA, 3rd, Loyd JE, et al. Heterozygous germline mutations in BMPR2, encoding a TGF-beta receptor, cause familial primary pulmonary hypertension. Nat Genet. 2000;26(1):81–4.

5. Evans JD, Girerd B, Montani D, Wang XJ, Galiè N, Austin ED, et al. BMPR2 mutations and survival in pulmonary arterial hypertension: an individual participant data meta-analysis. Lancet Respir Med. 2016;4(2):129–37.

6. Pullamsetti SS, Savai R, Janssen W, Dahal BK, Seeger W, Grimminger F, et al. Inflammation, immunological reaction and role of infection in pulmonary hypertension. Clin Microbiol Infect. 2011;17(1):7–14.

7. Price LC, Wort SJ, Perros F, Dorfmüller P, Huertas A, Montani D, et al. Inflammation in pulmonary arterial hypertension. Chest. 2012;141(1):210–21.

8. Rabinovitch M, Guignabert C, Humbert M, Nicolls MR. Inflammation and immunity in the pathogenesis of pulmonary arterial hypertension. Circ Res. 2014;115(1):165–75.

9. Soon E, Holmes AM, Treacy CM, Doughty NJ, Southgate L, Machado RD, et al. Elevated levels of inflammatory cytokines predict survival in idiopathic and familial pulmonary arterial hypertension. Circulation. 2010;122(9):920–7.

10. Gräf S, Haimel M, Bleda M, Hadinnapola C, Southgate L, Li W, et al. Identification of rare sequence variation underlying heritable pulmonary arterial hypertension. Nat Commun. 2018;9(1):1416.

11. Turro E, Astle WJ, Megy K, Gräf S, Greene D, Shamardina O, et al. Whole-genome sequencing of patients with rare diseases in a national health system. Nature. 2020;583(7814):96–102.

12. Hoeper MM, Bogaard HJ, Condliffe R, Frantz R, Khanna D, Kurzyna M, et al. Definitions and diagnosis of pulmonary hypertension. J Am Coll Cardiol. 2013;62(25 Suppl):D42–50.

13. Simonneau G, Gatzoulis MA, Adatia I, Celermajer D, Denton C, Ghofrani A, et al. Updated clinical classification of pulmonary hypertension. J Am Coll Cardiol. 2013;62(25 Suppl):D34–41.

14. Benjamini Y, Hochberg Y. Controlling the False Discovery Rate: A Practical and Powerful Approach to Multiple Testing. Journal of the Royal Statistical Society Series B (Methodological). 1995;57(1):289–300.

15. McLaughlin VV, Shillington A, Rich S. Survival in primary pulmonary hypertension: the impact of epoprostenol therapy. Circulation. 2002;106(12):1477–82.

16. Humbert M, Sitbon O, Chaouat A, Bertocchi M, Habib G, Gressin V, et al. Survival in patients with idiopathic, familial, and anorexigen-associated pulmonary arterial hypertension in the modern management era. Circulation. 2010;122(2):156–63.

17. Galiè N, Humbert M, Vachiery JL, Gibbs S, Lang I, Torbicki A, et al. 2015 ESC/ERS Guidelines for the Diagnosis and Treatment of Pulmonary Hypertension. Rev Esp Cardiol (Engl Ed). 2016;69(2):177.

18. Cracowski JL, Chabot F, Labarère J, Faure P, Degano B, Schwebel C, et al. Proinflammatory cytokine levels are linked to death in pulmonary arterial hypertension. Eur Respir J. 2014;43(3):915–7.

19. Simpson CE, Chen JY, Damico RL, Hassoun PM, Martin LJ, Yang J, et al. Cellular sources of interleukin-6 and associations with clinical phenotypes and outcomes in pulmonary arterial hypertension. Eur Respir J. 2020;55(4).

20. Heresi GA, Aytekin M, Hammel JP, Wang S, Chatterjee S, Dweik RA. Plasma interleukin-6 adds prognostic information in pulmonary arterial hypertension. Eur Respir J. 2014;43(3):912–4.

21. Humbert M, Monti G, Brenot F, Sitbon O, Portier A, Grangeot-Keros L, et al. Increased interleukin-1 and interleukin-6 serum concentrations in severe primary pulmonary hypertension. Am J Respir Crit Care Med. 1995;151(5):1628–31.

22. Tamura Y, Phan C, Tu L, Le Hiress M, Thuillet R, Jutant EM, et al. Ectopic upregulation of membrane-bound IL6R drives vascular remodeling in pulmonary arterial hypertension. J Clin Invest. 2018;128(5):1956–70.

23. Arita Y, Sakata Y, Sudo T, Maeda T, Matsuoka K, Tamai K, et al. The efficacy of tocilizumab in a patient with pulmonary arterial hypertension associated with Castleman’s disease. Heart Vessels. 2010;25(5):444–7.

24. Furuya Y, Satoh T, Kuwana M. Interleukin-6 as a potential therapeutic target for pulmonary arterial hypertension. Int J Rheumatol. 2010;2010:720305.

25. Kadavath S, Zapantis E, Zolty R, Efthimiou P. A novel therapeutic approach in pulmonary arterial hypertension as a complication of adult-onset Still’s disease: targeting IL-6. Int J Rheum Dis. 2014;17(3):336–40.

26. Taniguchi K, Shimazaki C, Fujimoto Y, Shimura K, Uchiyama H, Matsumoto Y, et al. Tocilizumab is effective for pulmonary hypertension associated with multicentric Castleman’s disease. Int J Hematol. 2009;90(1):99–102.

27. Soon E, Crosby A, Southwood M, Yang P, Tajsic T, Toshner M, et al. Bone morphogenetic protein receptor type II deficiency and increased inflammatory cytokine production. A gateway to pulmonary arterial hypertension. Am J Respir Crit Care Med. 2015;192(7):859–72.

28. Hurst LA, Dunmore BJ, Long L, Crosby A, Al-Lamki R, Deighton J, et al. TNFα drives pulmonary arterial hypertension by suppressing the BMP type-II receptor and altering NOTCH signalling. Nat Commun. 2017;8:14079.

