## Supplement for "Direct influence of BMPR2 mutations on cytokine patterns and biomarker effectiveness in pulmonary arterial hypertension"

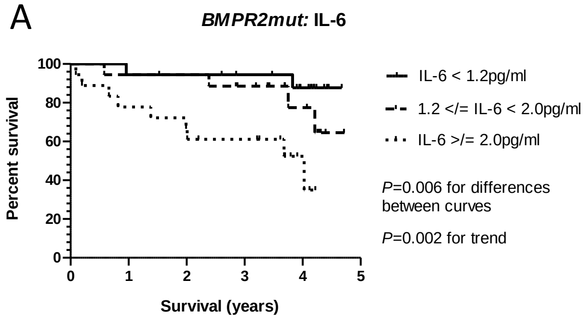


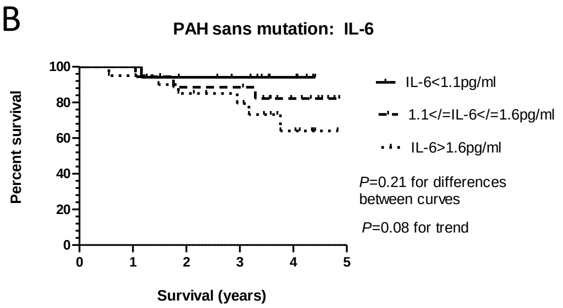


| Patients at risk | 0yrs | 1yr | 2yrs | 3yrs | 4yrs |
|---|---|---|---|---|---|
| BMPR2mut: IL-6 < 1.2 pg/ml | 18 | 18 | 18 | 16 | 11 |
| BMPR2mut: 1.2 </= IL-6 < 2.0 pg/ml | 18 | 18 | 17 | 14 | 08 |
| BMPR2mut: IL-6 >/= 2.0 pg/ml | 18 | 15 | 13 | 11 | 04 |

| Patients at risk | 0yrs | 1yr | 2yrs | 3yrs | 4yrs |
|---|---|---|---|---|---|
| PAH sans mutation: IL-6 < 1.2 pg/ml | 17 | 17 | 15 | 13 | 08 |
| PAH sans mutation: 1.2 </= IL-6 < 2.0 pg/ml | 18 | 18 | 16 | 16 | 09 |
| PAH sans mutation: IL-6 >/= 2.0 pg/ml | 20 | 20 | 18 | 16 | 08 |

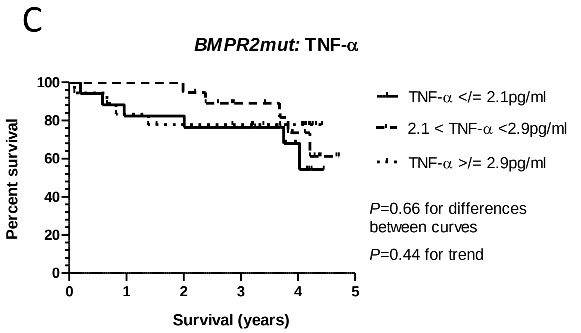


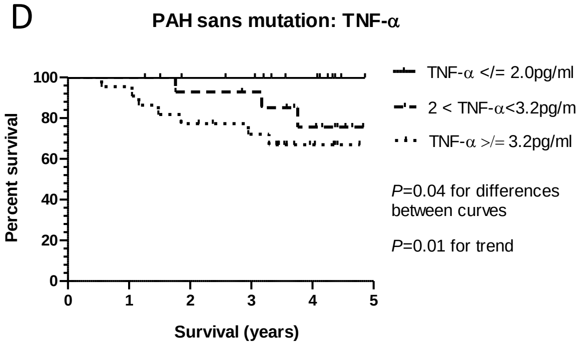


| Patients at risk | 0yrs | 1yr | 2yrs | 3yrs | 4yrs |
|---|---|---|---|---|---|
| BMPR2mut: TNF-α </= 2.1pg/ml | 17 | 15 | 15 | 14 | 06 |
| BMPR2mut: 2.1 < TNF-α < 2.9pg/ml | 19 | 19 | 19 | 14 | 08 |
| BMPR2mut: TNF-α > 2.9 pg/ml | 18 | 16 | 14 | 13 | 09 |

| Patients at risk | 0yrs | 1yr | 2yrs | 3yrs | 4yrs |
|---|---|---|---|---|---|
| PAH sans mutation: TNF-α </= 2.1pg/ml | 17 | 17 | 16 | 15 | 11 |
| PAH sans mutation: 2.1 < TNF-α < 2.9pg/ml | 15 | 15 | 14 | 13 | 09 |
| PAH sans mutation: TNF-α > 2.9 pg/ml | 22 | 22 | 18 | 15 | 06 |

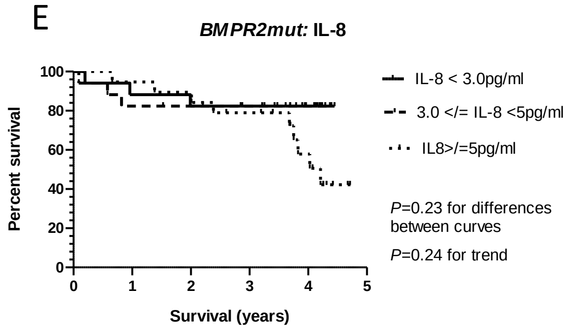


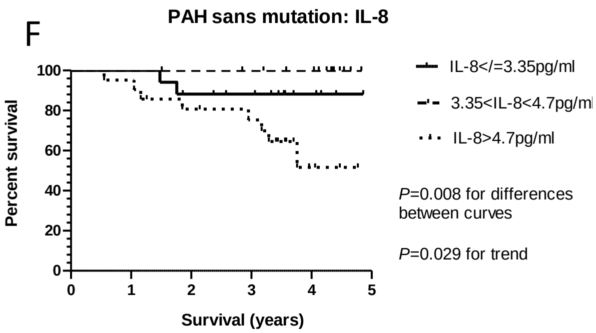


| Patients at risk | 0yrs | 1yr | 2yrs | 3yrs | 4yrs |
|---|---|---|---|---|---|
| BMPR2mut: IL-8 < 3.0 pg/ml | 17 | 16 | 15 | 12 | 05 |
| BMPR2mut: 3.0 </= IL-8 < 5.0 pg/ml | 17 | 15 | 14 | 13 | 09 |
| BMPR2mut: IL-8 >/= 5.0 pg/ml | 19 | 19 | 18 | 15 | 09 |

| Patients at risk | 0yrs | 1yr | 2yrs | 3yrs | 4yrs |
|---|---|---|---|---|---|
| PAH sans mutation: IL-8 < 3.0 pg/ml | 17 | 17 | 15 | 13 | 07 |
| PAH sans mutation: 3.0 </= IL-8 < 5.0 pg/ml | 16 | 16 | 16 | 15 | 13 |
| PAH sans mutation: IL-8 >/= 5.0 pg/ml | 21 | 21 | 17 | 15 | 04 |

**Supplementary Figure 1**

Kaplan-Meier analyses based on cytokine values. Graphs (A-B) show Kaplan-Meier curves based on tertiles of IL-6 values derived from the current cohort of *BMPR2mut* (A) and PAH sans mutation (B) patients. Graphs (C-D) show Kaplan-Meier curves based on tertiles of TNF-α values derived from the current cohort of *BMPR2mut* (C) and PAH sans mutation (D) patients. Graphs (E-F) show Kaplan-Meier curves based on tertiles of IL-8 values derived from the current cohort of *BMPR2mut* (E) and PAH sans mutation (F) patients.

A

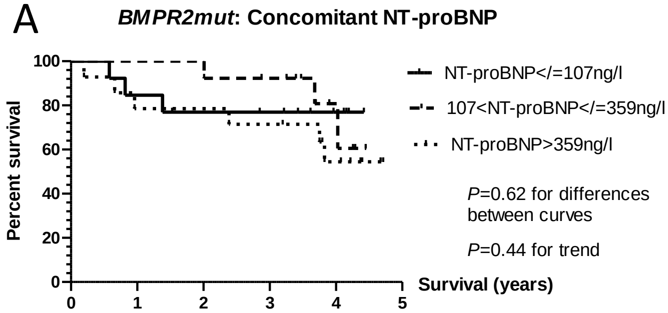


B

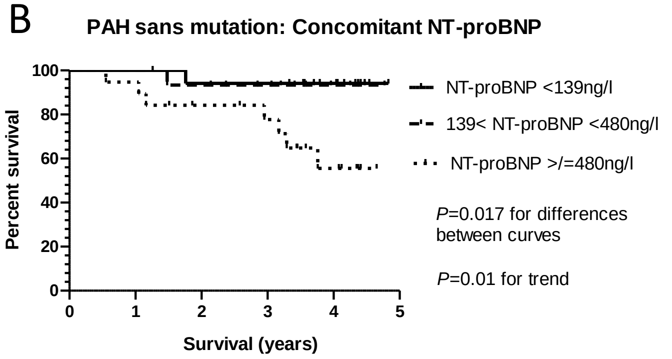


| Patients at risk | 0yrs | 1yr | 2yrs | 3yrs | 4yrs |
|---|---|---|---|---|---|
| BMPR2mut: NT-proBNP </= 107ng/l | 13 | 12 | 10 | 9 | 5 |
| BMPR2mut: 107 < NT-proBNP </= 359ng/l | 13 | 13 | 13 | 12 | 5 |
| BMPR2mut: NT-proBNP >359 ng/l | 14 | 13 | 12 | 11 | 7 |

| Patients at risk | 0yrs | 1yr | 2yrs | 3yrs | 4yrs |
|---|---|---|---|---|---|
| PAH sans mutation: NT-proBNP < 139ng/l | 17 | 17 | 17 | 17 | 10 |
| PAH sans mutation: 139 < NT-proBNP <480ng/l | 16 | 16 | 15 | 12 | 6 |
| PAH sans mutation: NT-proBNP >480 ng/l | 19 | 19 | 16 | 13 | 7 |

C

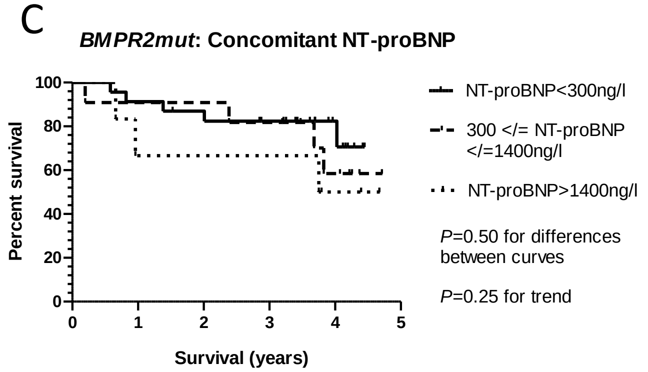


D

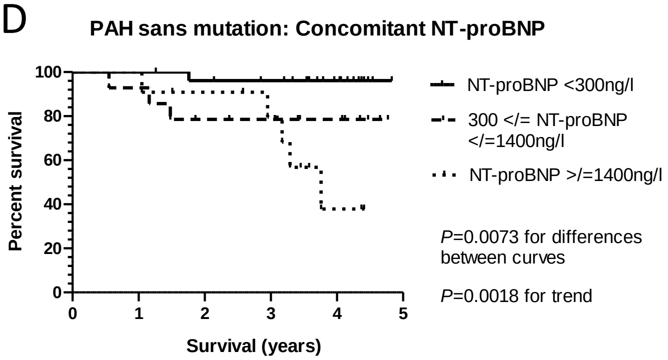


| Patients at risk | 0yrs | 1yr | 2yrs | 3yrs | 4yrs |
|---|---|---|---|---|---|
| BMPR2mut: NT-proBNP < 300ng/l | 23 | 22 | 20 | 17 | 8 |
| BMPR2mut: 300 < NT-proBNP </= 1400ng/l | 11 | 11 | 11 | 10 | 6 |
| BMPR2mut: NT-proBNP > 1400ng/l | 6 | 5 | 5 | 5 | 3 |

| Patients at risk | 0yrs | 1yr | 2yrs | 3yrs | 4yrs |
|---|---|---|---|---|---|
| PAH sans mutation: NT-proBNP < 300ng/l | 27 | 27 | 26 | 24 | 12 |
| PAH sans mutation : 300 < NT-proBNP </= 1400ng/l | 14 | 14 | 11 | 9 | 7 |
| PAH sans mutation: NT-proBNP > 1400ng/l | 11 | 11 | 10 | 8 | 3 |

**Supplementary Figure 2**

Kaplan-Meier analyses based on NT-proBNP values. Graphs (A-B) show Kaplan-Meier curves based on tertiles derived from the current cohort of *BMPR2mut* (A) and PAH sans mutation (B) patients. Graphs (C-D) show Kaplan-Meier curves based on European Respiratory Society definitions of ‘low’ (<300ng/l) and ‘high’ (>1,400ng/l) values of NT-proBNP for *BMPR2mut* (C) and PAH sans mutation (D) patients.

A

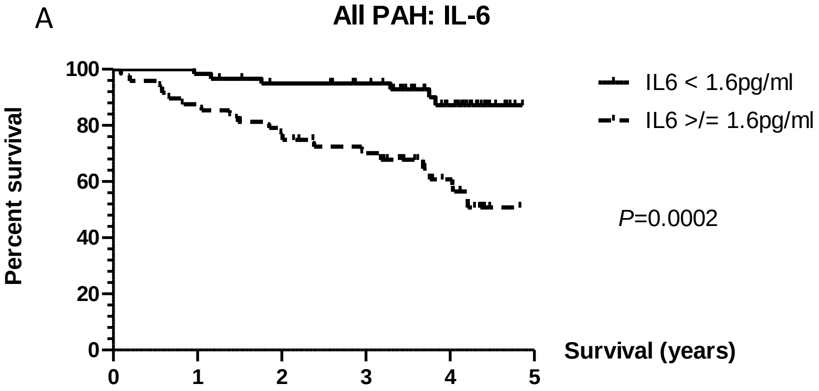


| Patients at risk | 0yrs | 1yr | 2yrs | 3yrs | 4yrs |
|---|---|---|---|---|---|
| All PAH: IL-6< 1.6pg/ml | 60 | 60 | 55 | 49 | 28 |
| All PAH: IL-6> 1.6pg/ml | 48 | 43 | 37 | 31 | 15 |

B

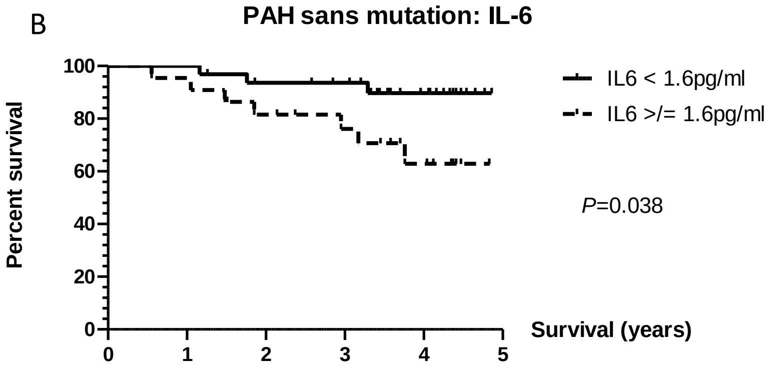


| Patients at risk | 0yrs | 1yr | 2yrs | 3yrs | 4yrs |
|---|---|---|---|---|---|
| PAH sans mutation: IL-6< 1.6pg/ml | 32 | 32 | 29 | 27 | 14 |
| PAH sans mutation: IL-6> 1.6pg/ml | 22 | 22 | 18 | 15 | 09 |

**Supplementary Figure 3**

Kaplan-Meier analyses based on a proposed cut-off value of 1.6pg/ml (median value for IL-6 in the *BMPR2mut* cohort and in the IPAH Simpson cohort). Graph (A) shows a Kaplan-Meier curve from the entire group of PAH patients and graph (B) shows the corresponding analysis in PAH without driving mutations alone.
